# Process evaluation of pragmatic cluster randomized trials of digital adherence technologies for tuberculosis treatment support: a mixed methods study in five countries

**DOI:** 10.1101/2024.11.26.24317987

**Authors:** N. Madden, A. Tadesse, A. Leung, B. Gonçalves Tasca, J. Alacapa, N. Deyanova, N. Ndlovu, N. Mokone, B Onjare, A. Mganga, K. van Kalmthout, D. Jerene, K. Fielding

## Abstract

**Background:** Digital adherence technologies (DATs) could improve the person-centredness of TB treatment. Acceptability of DATs is high, though evidence of their effectiveness is varied. Our objective was to understand the fidelity of DAT interventions within five cluster-randomized trials.

**Methods:** Two DATs (smart pill box, medication labels) were assessed, with real-time adherence data made available to healthcare providers (HCPs) on a digital platform in Ethiopia, the Philippines, South Africa, Tanzania, and Ukraine. The process evaluation framework assessed four components: inputs, processes, outputs, and outcomes. Fidelity of the most important intervention components was evaluated by quantitative indicators, with analysis conducted by country and DAT type. Content analysis of qualitative sub-studies supplemented some indicators.

**Results:** Engagement with DATs was high among persons with TB (PwTB). Pillbox users showed high levels of sustained engagement, with digitally recorded doses ranging from 82% to 91%. Sole ownership of a mobile phone was highest in South Africa (90%) and lowest in the Philippines (63%). Differences were also observed in the frequency of logins by HCPs to the adherence platform and the type of device used. In the Philippines and Ukraine, >50% of logins were from mobile phones. In Ethiopia, Tanzania, and Ukraine there was at least one login to the platform on 71% of weekdays per facility, compared with the Philippines and South Africa at 42% and 52%, respectively. A feeling of connection between PwTB and their HCP was reported by over 95% of participants surveyed in Ethiopia and Tanzania, this was 84% in South Africa and 76% Philippines, a finding underpinned by qualitative data.

**Conclusion:** We observed varying levels of intervention fidelity between countries. Timeliness and intensity of utilization of real-time data, and taking required actions are impacted by staff and health system capacity. Acceptance of DATs is high; therefore, future work should focus on identifying optimal intervention strategies.

## Introduction

Addressing the global challenge of tuberculosis requires concerted efforts that improve both treatment coverage and outcomes through person-centred approaches. Among other interventions, digital adherence technologies (DATs) have the potential to enhance the person-centredness of TB treatment and their effectiveness in improving treatment outcomes remain a subject of ongoing studies. Emerging data from recent studies provided mixed results, suggesting the need for more evidence in this area. (1), (2), (3), (4), (5), (6).

We conducted cluster randomized trials in five countries under the Adherence Support Coalition to End TB (ASCENT) project to fill this evidence gap (7), (8). The results from the five trials showed no difference in poor end of treatment outcome between the intervention and control arms among people with drug-sensitive TB (DS-TB) (9), (10). Findings from accompanying sub-studies showed high levels of acceptability of the DATs suggesting that DATs combined with differentiated care have direct benefits to persons with TB (PwTB), (11), (12), (13), (14), (15), (16).

Further scale-up of DATs should therefore be supported by a thorough evaluation of the process of implementation, understanding both context and process of implementing the intervention being studied (17). Process evaluation is a tool to assess fidelity and quality of implementation, clarify causal mechanisms, and identify contextual factors associated with various outcomes. Process evaluations provide critical information to enhance understanding of findings from pragmatic randomized controlled trials (RCTs) (18). Understanding implementation is particularly important for pragmatic trials, as onsite partners rather than research teams deliver the intervention (19). Both the content of an intervention and how it is delivered can be amended by those applying the intervention, therefore, establishing fidelity to the intervention is key to understanding reasons an intervention succeeded or failed, particularly for complex interventions such as DATs (20). We aimed to understand the fidelity of the intervention under the five pragmatic trials of DATs among adults with DS-TB (7), (8).

## Methods

### Design

We conducted independent, pragmatic, cluster-randomised trials in the Philippines, Ethiopia, South Africa, Tanzania, and Ukraine to assess whether digital adherence technologies coupled with support for those with an indication of poor medication adherence can improve treatment outcomes among adults with DS-TB. The trial protocols are described in detail elsewhere (7), (8), (21). In brief, the trials fall closer to the pragmatic side of the pragmatic/explanatory continuum based on PwTB populations, delivery of the intervention and trial outcomes (22). Two DAT types, smart pillboxes and medication labels, were evaluated.

### DAT interventions

PwTB using the pillbox received daily audio or audio-visual reminders to take their treatment. For the medication labels, PwTB were expected to send a free SMS to a shortcode daily, when treatment was taken, with a code on the medication label. Pillbox opening and receipt of SMS from PwTB, as proxies for doses taken, were captured digitally onto the adherence platform. If there was no recording of a dose taken at a country-specific predefined time, an automated SMS reminder to take treatment was sent, on the day, and the following day in the continued absence of a box-opening by the PwTB or SMS. No reminders were sent in Ukraine based on the decision from the Ukrainian research team. In the other four countries, the participant could opt out of the reminder messages that follow when a dose was not confirmed by a set time each day, though information on which participants opted out was not available. South Africa experienced some issues with sending of multiple reminder messages to participants who had taken their dose. It is possible that during some periods of implementation in South Africa the reminder message was switched off due to this technical issue.

The recording of doses taken on the adherence platform allowed individual-level DAT engagement information to be generated. The TB HCP was expected to review these data regularly and initiate differentiated care based on pre-defined adherence patterns. The adherence platform records, for each PwTB, a treatment-day as either: digitally recorded (pillbox opened on the day, or SMS sent); manually recorded (no recording that the pillbox was opened or SMS sent but the HCP has confirmed a dose was taken by contacting the PwTB); dose missed (no recording that the pillbox was opened or SMS sent and the HCP has confirmed a dose was not taken by calling the PwTB); and unknown (no recording that the pillbox was opened or SMS sent and no additional information from the HCP). HCP confirmation that a dose was or was not taken could take place at any time on or after the treatment-day.

### Context for the intervention

The infrastructure to deliver the intervention, training of HCPs, and support visits to health facilities made up the resources to support the implementation of the intervention. The infrastructure included the online adherence platform which both DATs were linked to, hosting of the platform in each country, hardware (tablets, desktops) and data services for HCPs to deliver the intervention. Training of HCPs implementing the intervention included formal training sessions and mentoring on the effective utilization of the adherence platform, PwTB monitoring, manual dosing procedures, and the provision of differentiated care. Training sessions were repeated, including for newly recruited staff. Additionally, HCPs from departments other than TB, who did not receive formal training, but were added to the cohort of staff who could initiate a participant on a DAT, received cascade and on-the-job training from the TB focal person who was formally trained previously. This style of training was utilized when core TB staff were redeployed to the COVID-19 response, or when capacity was low.

The study team trained HCPs on DATs, provided implementation support, and addressed technical issues with DATs on the adherence platform. In the preparatory phase that took place 3-6 months before the trial enrollment phase, the focus included facility readiness to implement the intervention and operationalizing the use of DATs. Stock levels of DATs were monitored during visits, as well as recording of previous stockouts. Defective or malfunctioning DATs were noted and replaced.

### Process evaluation framework

A process evaluation framework provides understanding of why and how the intervention was able to lead to improved treatment outcomes or suggests reasons for the lack of any observed change. Our framework was based on four components: inputs, processes, outputs, and outcomes (Figure 2, supplementary material). Quantitative indicators focused on the reach and fidelity of the intervention, complemented by qualitative indicators, Table 1 summarizes key indicators.

**Table 1:** Summary of process evaluation indicators and their data sources.

|  | Indicators | Data Sources | Content Analysis categorization |
| --- | --- | --- | --- |
| Input |  |  |  |
| 1 | No. of Healthcare providers received implementation training during the pilot and main enrolment period. | Country training report | HCPs implementation training |
| 2 | No. of implementation support visits made to health facilities by ASCENT staff during the main enrolment period. | ASCENT facility visit – call Log | NA |
| 3 | Percentage of PwTB who own a mobile phone which is not shared | ASCENT Sub-study 1 | PwTB with access to or that own a mobile phone |
| Process |  |  |  |
| 4 | No. of automated SMS reminders sent by adherence platform per PwTB enrolled during the main enrolment period. | Adherence platform SMS Log | Automated SMS reminders sent by adherence platform |
| 5 | Percentage of treatment days with an automated SMS reminder sent by the adherence platform | Adherence platform SMS Log |  |
| 6 | No./ Percentage of PwTB that switch from medication label to smart pillbox during their treatment<br>No./ Percentage of PwTB in labels arm that initiated on pillbox at the start of their treatment | Adherence platform PwTB record | PwTB switch from medication labels to smart pill box during treatment |
| 7 | No. of home visits per PwTB by initial DAT received during the main enrolment period. | Adherence platform HCWs action report | Home visits during the main enrolment period |
| 8 | No./ Percentage of PwTB who were shown their adherence data at the facility by the HCP | ASCENT Sub-study 1 | NA |
| 9 | Total number of adherence platform logins per facility during main enrolment period | Adherence platform usage record | NA |
| 10 | Percentage of weekdays in the main enrolment period where there was at least one visit to the adherence platform at facility level | Adherence platform usage record | HCPs interaction with the adherence platform |
| 11 | Average minutes healthcare worker spent on platform per day | Adherence platform usage record |  |
| 12 | Average percentage of ASCENT tablet use to access adherence platform during main enrolment period. | Adherence platform usage record |  |
| Output |  |  |  |
| 13 | Percentage of PwTB in who started a DAT in the main enrollment period | Adherence platform PwTB record & facility TB register | NA |
| 14 | Average percentage of doses digitally recorded of all doses recorded during main enrolment period. | Adherence platform dosing report | NA |
| 15 | Patterns of manual doses added to the adherence platform more than 7 days after the original date during main enrolment period. | Adherence platform dosing report | NA |
| Outcome |  |  |  |
| 16 | Can the use of DATs and differentiated care influence PwTB-HCPs relationship? | ASCENT Sub-study 1, 2 and 3 | People with TB – HCP relationship |

Sub-study 1: cross-sectional survey to assess the acceptability and feasibility of the interventions for people with TB (conducted in all countries except Ukraine); Sub-study 2: in-depth interviews of people with TB to assess the acceptability and feasibility of DATs and differentiated care (conducted in all countries except Ukraine); Sub-study 3: in-depth interviews of healthcare workers and key stakeholders' perspectives to assess the acceptability and feasibility of DATs and differentiated care (conducted in all countries). NA not applicable; PwTB person with TB; SMS short message service; DAT digital adherence technology; HCP healthcare provider

### Data sources

We used multiple data sources for the process evaluation: the adherence platform (dosing, support actions and engagement, and SMS logs); data from training logs and implementation support visits; and data from related sub-studies (11), (13), (14), (15), (16) where relevant. Data were collected from 1 December 2020 to 31 August 2022 covering the full period of recruitment of participants to the effectiveness trial. Written informed consents of PwTB for the main effectiveness study were collected from PwTB by the TB care providers at the intervention facilities. Prior to participation in qualitative interviews, written informed consent was also obtained. In addition, a waiver of consent was obtained to access TB register data. See additional methods in the supplement section 1.1 to 1.3, and Table 1.

### Analysis

We used descriptive analysis stratified by country and DAT type using R and Stata 18, to provide an overview of process evaluation indicators. For platform logins, the percentage of weekdays in the main enrolment period where there was at least one visit to the adherence platform at each facility was calculated. For SMS reminder indicators, the number of treatment days was calculated for each participant. DAT engagement indicators were represented by digitally recorded doses by the participants, and manually added doses by the HCP. Participants were analyzed primarily based on the DAT they initiated treatment with.

We restricted platform data to the trial main enrolment phase which ranged from 12-13 months, from June 2021-August 2023, and the associated 6-month follow-up period whilst PwTB were on treatment. We extracted the platform data in June 2023. We conducted a content analysis of the two qualitative sub-studies (among PwTB, and HCP and stakeholders), guided by the process evaluation indicators. Indicators were grouped by thematic similarity to identify related content in each qualitative sub-study, as reported in table 1.

## Results

The DAT intervention was implemented in 162 facilities across the five countries, enrolling 10,377 participants starting a DAT in the intervention arms.

### Input indicators

The average number of healthcare providers trained per facility ranged from 2 and 7 across the five countries (Table 2). Training before the intervention initiation was a condition for proper DAT implementation in healthcare facilities. HCPs noted that staff shortages caused by COVID-19 impeded the effective multiplication of training by peers who received implementation training. In the Philippines, HCPs mentioned wanting more training sessions and staff shortages making it difficult to properly instruct colleagues.

**Table 2:** Results of indicators assessing implementation inputs by country and DAT type (where relevant)

|  | Indicator | Ethiopia | The Philippines | South Africa | Tanzania | Ukraine |
| --- | --- | --- | --- | --- | --- | --- |
| 1 | Number of HCPs trained/facility<sup>a</sup> | 3.0 (155/52) | 4.9 (157/32) | 2.6 (79/30) | 2.0 (72/36) | 7.4(89/12) |
| 2 | Number of implementation support visits/facility<sup>b</sup> | 1.7 (89/52) | 2.4 (78/32) | 1.3 (39/30) | 1.8 (65/36) | 0.7 (8/12) |
| 3 | Phone ownership (not shared) - among PwTB using the pillbox<sup>c</sup> | 72% (36/50) | 62% (31/50) | 90% (44/49) | 70% (43/61) | NA |
|  | Phone ownership (not shared) – among PwTB using the labels<sup>c</sup> | 70% (35/50) | 64% (33/52) | 97% (28/29) | 68% (26/38) | NA |

HCP health care provider; PwTB person with TB; NA not applicable.
<sup>a</sup> training log; <sup>b</sup> implementation support visit log; <sup>c</sup> data from sub-study 1.

*“I wish there would be another training so that I can really know the program. I was just able to do that by fiddling with my cellphone.”* – HCP in the Philippines.

Cellphone ownership (not shared) among PwTB was highest in South Africa (90% pillbox, 97% medication labels) and lowest in the Philippines (63% -pillbox, 64% medication labels). In Ethiopia and Tanzania, it was similar across DAT types, ranging from 68% to 72%. For the implementation of medication labels, having access to a cellphone was a condition to be offered a DAT. The expansion of mobile coverage and cellphone usage in Ethiopia was reported as a facilitating factor for DAT usage. It was not uncommon for people to share a cellphone with other family members. People would stop using the medication labels or change to a pillbox if their cellphone was damaged or lost.

*“That’s when we have a problem when the patient doesn’t have a cell phone and they don’t have a support system, so we give them a pill box.”*-HCP in the Philippines.

### Process indicators

Process indicators are summarized in table 3. The total SMS reminders sent per PwTB was highest in South Africa and Ethiopia (60.7 and 53.1/PwTB) and lowest in Tanzania (30.9/participant). The percentage of days on treatment during which participants received same-day reminders was higher among participants using labels versus pillboxes in Ethiopia (32% vs 23%), the Philippines (38% versus 18%) and South Africa (37% versus 21%), and similar for Tanzania (15% versus 13%). Reminders sent for a previous day’s missed dose showed similar differences by DAT type for the Philippines and South Africa.

**Table 3:**
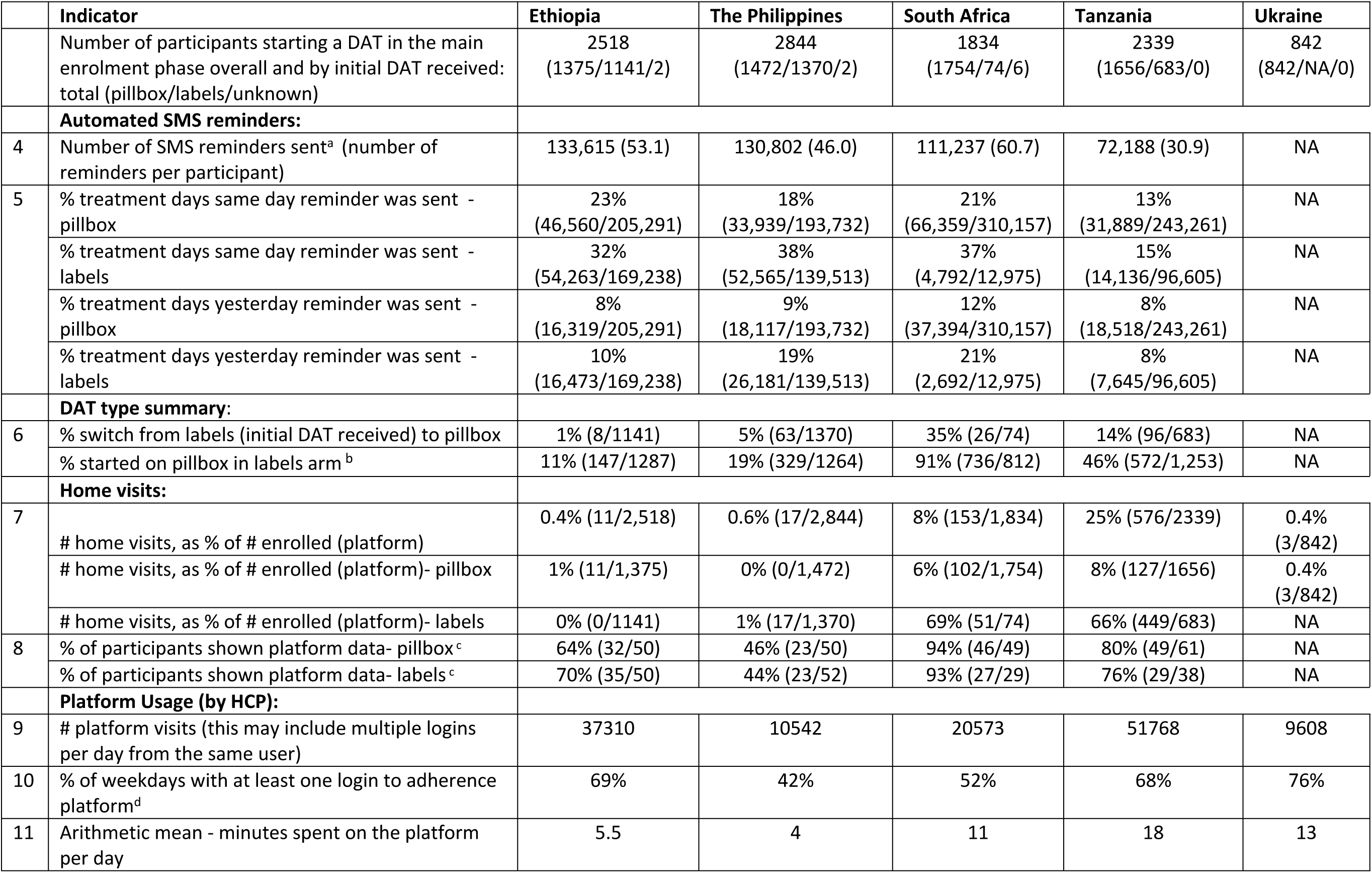
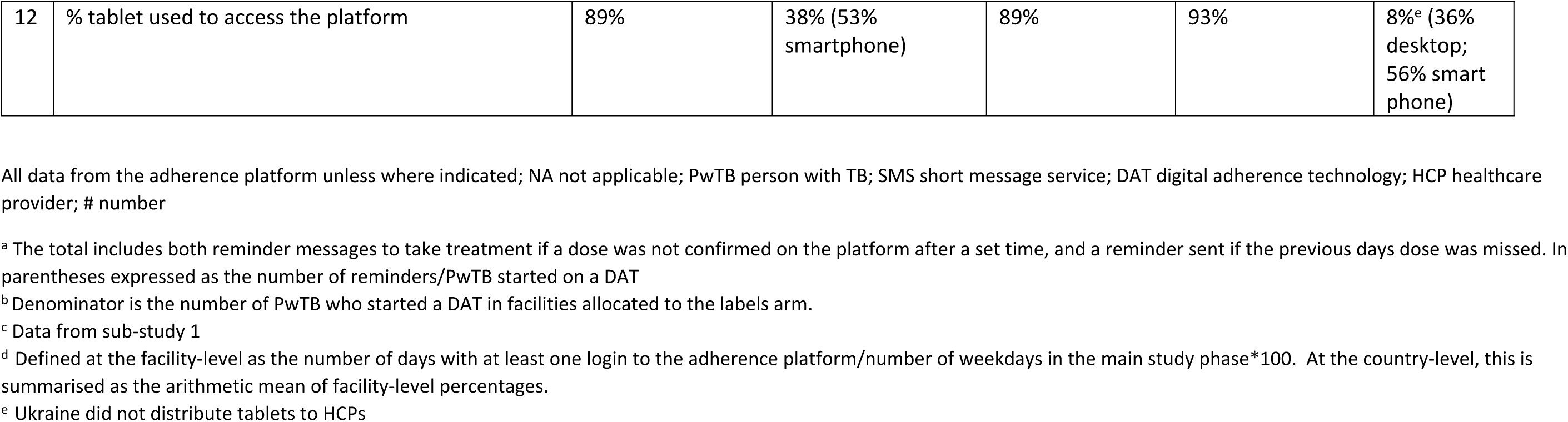
Results of indicators assessing implementation processes by country and DAT type (where relevant)

|  | Indicator | Ethiopia | The Philippines | South Africa | Tanzania | Ukraine |
| --- | --- | --- | --- | --- | --- | --- |
|  | Number of participants starting a DAT in the main enrolment phase overall and by initial DAT received: total (pillbox/labels/unknown) | 2518<br>(1375/1141/2) | 2844<br>(1472/1370/2) | 1834<br>(1754/74/6) | 2339<br>(1656/683/0) | 842<br>(842/NA/0) |
|  | **Automated SMS reminders:** |  |  |  |  |  |
| 4 | Number of SMS reminders sent<sup>a</sup> (number of reminders per participant) | 133,615 (53.1) | 130,802 (46.0) | 111,237 (60.7) | 72,188 (30.9) | NA |
| 5 | % treatment days same day reminder was sent - pillbox | 23%<br>(46,560/205,291) | 18%<br>(33,939/193,732) | 21%<br>(66,359/310,157) | 13%<br>(31,889/243,261) | NA |
|  | % treatment days same day reminder was sent - labels | 32%<br>(54,263/169,238) | 38%<br>(52,565/139,513) | 37%<br>(4,792/12,975) | 15%<br>(14,136/96,605) | NA |
|  | % treatment days yesterday reminder was sent - pillbox | 8%<br>(16,319/205,291) | 9%<br>(18,117/193,732) | 12%<br>(37,394/310,157) | 8%<br>(18,518/243,261) | NA |
|  | % treatment days yesterday reminder was sent - labels | 10%<br>(16,473/169,238) | 19%<br>(26,181/139,513) | 21%<br>(2,692/12,975) | 8%<br>(7,645/96,605) | NA |
|  | **DAT type summary:** |  |  |  |  |  |
| 6 | % switch from labels (initial DAT received) to pillbox | 1% (8/1141) | 5% (63/1370) | 35% (26/74) | 14% (96/683) | NA |
|  | % started on pillbox in labels arm<sup>b</sup> | 11% (147/1287) | 19% (329/1264) | 91% (736/812) | 46% (572/1,253) | NA |
|  | **Home visits:** |  |  |  |  |  |
| 7 | # home visits, as % of # enrolled (platform) | 0.4% (11/2,518) | 0.6% (17/2,844) | 8% (153/1,834) | 25% (576/2339) | 0.4%<br>(3/842) |
|  | # home visits, as % of # enrolled (platform)- pillbox | 1% (11/1,375) | 0% (0/1,472) | 6% (102/1,754) | 8% (127/1656) | 0.4%<br>(3/842) |
|  | # home visits, as % of # enrolled (platform)- labels | 0% (0/1141) | 1% (17/1,370) | 69% (51/74) | 66% (449/683) | NA |
| 8 | % of participants shown platform data- pillbox<sup>c</sup> | 64% (32/50) | 46% (23/50) | 94% (46/49) | 80% (49/61) | NA |
|  | % of participants shown platform data- labels<sup>c</sup> | 70% (35/50) | 44% (23/52) | 93% (27/29) | 76% (29/38) | NA |
|  | **Platform Usage (by HCP):** |  |  |  |  |  |
| 9 | # platform visits (this may include multiple logins per day from the same user) | 37310 | 10542 | 20573 | 51768 | 9608 |
| 10 | % of weekdays with at least one login to adherence platform<sup>d</sup> | 69% | 42% | 52% | 68% | 76% |
| 11 | Arithmetic mean - minutes spent on the platform per day | 5.5 | 4 | 11 | 18 | 13 |
| 12 | % tablet used to access the platform | 89% | 38% (53%<br>smartphone) | 89% | 93% | 8%<sup>e</sup> (36%<br>desktop;<br>56% smart<br>phone) |

All data from the adherence platform unless where indicated; NA not applicable; PwTB person with TB; SMS short message service; DAT digital adherence technology; HCP healthcare provider; # number
<sup>a</sup> The total includes both reminder messages to take treatment if a dose was not confirmed on the platform after a set time, and a reminder sent if the previous days dose was missed. In parentheses expressed as the number of reminders/PwTB started on a DAT
<sup>b</sup> Denominator is the number of PwTB who started a DAT in facilities allocated to the labels arm.
<sup>c</sup> Data from sub-study 1
<sup>d</sup> Defined at the facility-level as the number of days with at least one login to the adherence platform/number of weekdays in the main study phase\*100. At the country-level, this is summarised as the arithmetic mean of facility-level percentages.
<sup>e</sup> Ukraine did not distribute tablets to HCPs

Individual reasons prompting reminder messages from the platform included forgetting to send a message when a dose was taken or forgetting to take the dose before the predefined time. Other factors prompting reminder messages were inability to read, technology fatigue, lack of understanding of DATs usage/purpose and low technology savviness. The latter was noted among the elderly population interviewed in particular.

Intermittent access to cell phones and needing to have a positive airtime balance to send messages also influenced the ability to send messages as instructed. Technical issues such as pillboxes running out of battery, unstable network and electricity supply also played a role in medication intake information not being registered timely on the platform. As a result, in some instances the adherence platform may have sent reminder messages even when medication was taken DATs utilized correctly.

*“Sometimes I receive a message that says, “you did not take your medicine today”. At that time, I came here [to the health facility] and explained that I have taken the medicine, but my house has a network problem. This inconvenience happens because of network, not because I didn’t take it”.* - PwTB in Ethiopia.

Among participants who initially received the labels, the percentage who switched to the pillbox varied from 1% (8/1141) in Ethopia to 35% (26/74) in South Africa. Among individuals enrolled from facilities randomised to the labels arm, 11% of participants started treatment using a pillbox in Ethiopia, while in the Philippines, Tanzania, and South Africa this percentage was 19%, 46% and 91% respectively. In South Africa, the labels intervention was stopped early due to multiple problems with implementing the SMS component and the facilities transitioned to the pillbox intervention (see supplementary information section 4.1).

Participants switching from medication labels to smart pillboxes, were mentioned in the Philippines, Tanzania and South Africa qualitative studies. Inability to read, technology fatigue, lack of familiarity with cellphones and forgetting to send an SMS were some of the reasons that influenced individuals to switch DATs. Technical issues such as technical glitches, leading to excessive SMS reminders received, or not being able to send the SMS code also influenced the decision to switch from the medication labels to the smart pill box. Cellphone damage or loss, the requirement of having positive airtime balance, recurrent power cuts and poor network connection were also mentioned as drivers of switching DATs.

*“I didn’t use the stickers [medication labels] during that time because I had lost my phone, do you get me? I arrived there and told the sister [TB Nurse], that’s when they gave me the box.”* - PwTB in South Africa.

Tanzania had the highest number of home visits per participant using a DAT recorded on the adherence platform at 25% (576/2339), this differed by DAT type, 8% (127/1656) for pillbox and 66% (449/683) for medication-label. In South Africa, 8% (153/1834) of participants had a home visit, 69% (51/74) of those using the labels, and 6% (102/1754) using the pillbox. The percentage of participants who had home visits in Ethiopia, the Philippines and Ukraine was <=1%.

Qualitative data indicates that in all countries other than Tanzania, staff shortages limited the number of home visits conducted. In Ukraine, home visits were not frequent due to the healthcare reform that led to staff reduction in specialized TB facilities, and displacement of both PwTB and HCP due to the war. The practice of home visits in Tanzania, was supported by a large cadre of community health workers, and they were part of routine care prior to the implementation of the intervention. Stakeholders’ interviews indicated that stigma-related issues influenced the feasibility of home visits. It was reported that some individuals would deliberately provide wrong addresses to avoid receiving visits.

*“Patients do not want to be visited to their home as there are TB patients whose families do not know that they have TB”* - HCP in Ethiopia.

PwTB experiencing homelessness, those not having a fixed address, relocations, or living in informal settings without an official address, were also barriers to HCP to providing home visits.

The percentage of participants who reported being shown adherence data by their provider in the feasibility and acceptability assessment ranged from 46% in the Philippines to 94% in South Africa in the pillbox arm. In the labels arm, this ranged from 44% in the Philippines to 93% in South Africa.

During the main enrolment phase, the number of HCP logins to the adherence platform ranged from 9,608 in Ukraine to 51,768 in Tanzania, while users in Ethiopia, the Philippines and South Africa logged in 37,310, 10,542 and 20, 573 times respectively (Table 3). The percentage of weekdays with at least one login to the adherence platform per facility ranged from 42% in the Philippines to 76% in Ukraine. It was 52% in South Africa, 68% in Tanzania and 69% in Ethiopia (figure 1). The mean number of minutes per facility per day spent on the platform was highest in Tanzania at 18 minutes, and lowest in the Philippines at 4 minutes.

**Figure 1:**
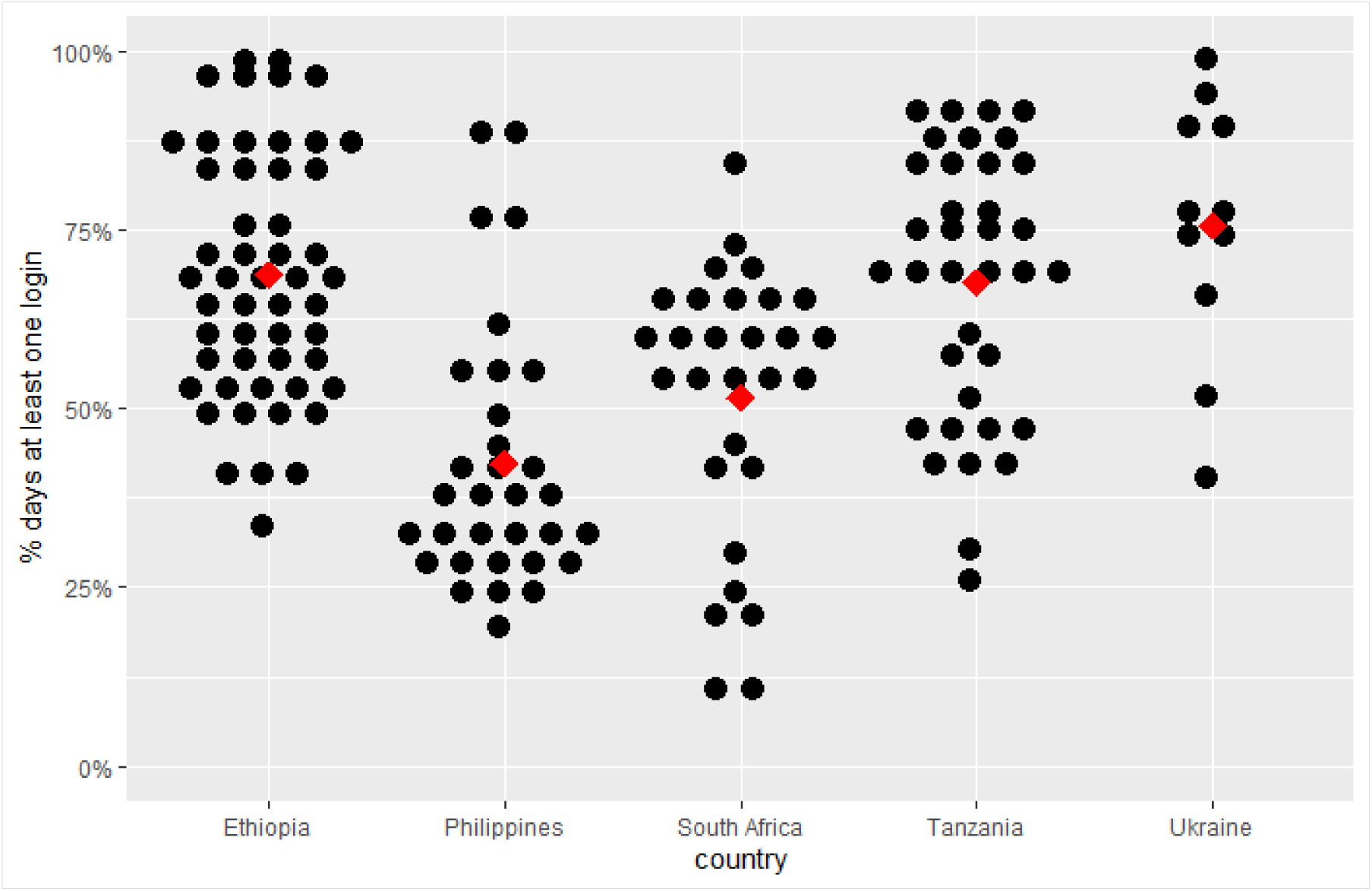
Percentage weekdays with at least one login to the adherence platform per facility. Each black dot represents a facility, the arithmetic mean is indicated by the red diamond.

HCPs from the Philippines, South Africa and Tanzania reported that the adherence platform reduced their workload and simplified activities. They were able to monitor multiple user’s treatment adherence at once.

*“This new system (adherence platform) has simplified our work, for instance when you enter the office in the morning, you look on the tablet to monitor patients’ treatment adherence, make follow up on patients’ with bad adherence.”* - HCP in Tanzania.

However, some HCPs also perceived that the platform added to their workload. In the Philippines, the adherence platform and the local Integrated Tuberculosis Information System (ITIS) operated simultaneously, requiring HCPs to input similar users’ information into both systems, increasing workload. In Ukraine, HCPs considered the process of validating dosing records burdensome.

*“But look, if the program (adherence platform) already recorded that the dose is missed, why does it have to be colored (verified) again? It is logical, right?”* -HCP in Ukraine.

Three types of devices were used to access the adherence platform; a tablet (provided by the ASCENT project), desktop or laptop computer or a smartphone. In Ethiopia and Tanzania, 89% of logins to the platform were from a tablet, it was 93% in South Africa. In the Philippines and Ukraine, most logins were from smartphones.

### Output and outcome indicators

Output and outcome indicators are summarized in table 4. The percentage of PwTB starting a DAT ranged from 55.2% in Ukraine to 73.5% in South Africa. The pillbox arm had both the lowest DAT coverage at 51.4% in the Philippines, and the highest at 79.8% in South Africa. Digitally recorded doses were higher in the smart pillbox versus the labels arms. The percentage of total doses digitally recorded ranged from 82% in Ukraine to 91% in Tanzania among pillbox users and 62% in South Africa and 84% in Tanzania among label users. The Philippines had the highest percentage of manual doses added to the adherence platform more than 7 days after the dose-day, when a dose was missed, in both pillbox and labels arm at 61%, and 55% respectively.

**Table 4:** Results of indicators assessing implementation outputs and outcomes by country and DAT type (where relevant)

|  | Indicator | Ethiopia | The Philippines | South Africa | Tanzania | Ukraine |
| --- | --- | --- | --- | --- | --- | --- |
|  | **Output:** |  |  |  |  |  |
|  | DAT Coverage |  |  |  |  |  |
| 13 | % of PwTB who started a DAT in the main enrolment period | NA<sup>a</sup> | 61.8%<br>(2,844/4,604) | 73.5%<br>(1,834/2,494) | 66.0%<br>(2,339/3,546) | 55.2%<br>(842/1,526) |
|  | % of PwTB who started a DAT in the main enrolment period - pillbox | NA<sup>a</sup> | 51.4%<br>(1,150/2,236) | 79.8%<br>(1,022/1,281) | 61.1%<br>(1,086/1,778) | 55.2%<br>(842/1,526) |
|  | % of PwTB who started a DAT in the main enrolment period - labels | NA<sup>a</sup> | 71.5%<br>(1,694/2,368) | 66.9%<br>(812/1,213) | 70.9%<br>(1,253/1,768) | NA |
|  | Number of PwTB - pillbox | 1375 | 1472 | 1754 | 1656 | 842 |
| 14 | % digital recorded doses-pillbox | 90%<br>(177,599/196,352) | 83%<br>(172,234/208,130) | 88%<br>(207,569/235,417) | 91%<br>(202,282/222,076) | 82%<br>(91,712/111,901) |
| 15 | % of manual doses added >7days after the dose-day – pillbox | 30% (5,075/16,845) | 61% (14,708/24,015) | 44% (3,966/9,075) | 32% (4,623/14,561) | 52% (6,858/13,206) |
|  | Number of PwTB - labels | 1141 | 1370 | 74 | 683 | - |
| 14 | % digital recorded doses-labels | 81%<br>(126,718/156,832) | 69%<br>(119,304/171,786) | 62% (3,815/6,154) | 84%<br>(63,862/76,231) | NA |
| 15 | % of manual doses added >7days after the dose-day – labels | 21% (6,016/28,782) | 55% (20,832/37,919) | 47% (541/1,149) | 26% (2,906/11,108) | NA |
|  | **Outcome:** |  |  |  |  |  |
| 16 | % participants - agreed that using DAT made them feel more connected to their HCPs – pillbox<sup>b</sup> | 100% (50/50) | 84.0% (42/50) | 91.8% (41/49) | 96.7% (59/61) | NA |
|  | % participants - agreed that using DAT made them feel more connected to their HCPs – labels<sup>b</sup> | 98.0% (49/50) | 69.2% (36/52) | 86.2% (25/29) | 94.7% (36/38) | NA |

All data from the adherence platform unless, where indicated
<sup>a</sup> Not applicable for the Ethiopian CRT as the study design differed. Data were only collected on people with TB who consented to the study <sup>b</sup> Data from Sub-study 1
DAT type missing for 10 individuals (2 The Philippines; 6 South Africa; and 2 Ethiopia). NA not applicable; PwTB person with TB; DAT digital adherence technology; HCP healthcare provider

Over 96% of sub study 1 survey respondents in Ethiopia and Tanzania agreed that using a pillbox DAT made them feel more connected to their HCP. In the Philippines, 70% of labels users reported better connection to their HCP, while this was over 95% in Ethiopia and Tanzania, with 86% of labels user in South Africa agreeing.

The feasibility and acceptability assessment revealed that HCPs perceived that the presence of DATs improved their relationships with adherent PwTB, by facilitating communication. According to HCPs, the adherence platform assisted them to act when they observed a PwTB not engaging with the DAT, thereby strengthening communication between the two parties. Some trust issues have also emerged mediated by the DAT technology. HCPs expressed scepticism on the proper usage of the DATs, they believed that certain PwTB were using the DATs without taking their medication to avoid being contacted by HCPs, which negatively impacted their relationship.

“*Well, it’s clear that if you are a person having a smart pill box and the doctor calls you 10 times in a month, you’ve already opened the smart pill box so that the doctor doesn’t bother you, doesn’t call you. And we have some (meaning patients) who say to us: you are so concerned about my health… Because we call them often*” - HCP in Ukraine.

## Discussion

We examined the fidelity of the implementation process and coverage of DATs and associated differentiated support under pragmatic trial conditions among adults with drug-sensitive TB in five countries. Results differ by country and provide insights into how the implementation of digital support mechanisms can be optimized in the future. Key findings indicate firstly that phone ownership was high overall. Secondly, engagement with DATs was high among participants, particularly in the pillbox arm, with digitally recorded doses ranging from 82% to 91%, with strong concurrence from the qualitative interviews. Thirdly, there was considerable variation in both method and frequency of HCPs accessing the digital adherence platform, with more than 50% of logins to the platform being from mobile phones rather than tablets or desktops in Ukraine and the Philippines. Additionally, the percentage of manual doses added to the adherence platform more than 7 days after the dose-day varied considerably by country, with the Philippines (61%), more than double Ethiopia (30%), in the pillbox arm.

The findings confirm the pragmatic nature of the trials with each country adapting their interventions to their local context. Moreover, this study provides a rich diversity of context for the implementation of DATs in terms of geographical, cultural, socio-economic, and demographic factors. Key findings indicate that integration of digital systems into daily workflow is complex, and the ability of health systems to adopt the new technology varies per country and facility. Health system and HCPs capacity has an impact on the ability to utilize real-time data. Reviewing adherence data daily may not be possible for all HCPs or necessary for all PwTB, and value may still be derived from less regular assessment of adherence and taking action where doses are missed, or prioritising adherence monitoring of certain population. DATs are a highly acceptable form of treatment support for PwTB and for the HCPs who can integrate digital systems with their regular tasks.

Access to a mobile phone was a requirement for using the medication label and the intervention can be implemented with a shared phone, although unintended disclosure of TB status is less likely on a personal phone. In an earlier study conducted in Tanzania, it was found that those with the highest engagement with 99DOTS were those who owned a mobile phone that was not shared (23). Access to mobile technology is key to the implementation of digital health interventions (24). Timely delivery of differentiated care is facilitated by a phone that is not shared. Automated SMS reminders when a digital dose was not received by the platform before a certain time each day was common but with variation between countries and more markedly DAT type. Qualitative findings on reasons to miss a dose, therefore, prompting a reminder message was aligned with other studies, where, the ability to read, technology fatigue and unstable network were cited as barriers to digital recording of a dose (25), (26), (27), (28).

Facility staff received formal training on the implementation of DATs and use of the adherence platform, varying from two HCPs per facility in Tanzania to seven per rayon in Ukraine. However, the cadre of staff trained varied. In Ukraine TB doctors would enroll PwTB on DATs, while in the Philippines, due to redeployment to COVID-19 services, additional staff from other disciplines received on the job training from TB focal persons enabling them to enroll and monitor PwTB. The lack of more comprehensive training of the staff providing secondary support in the Philippines may have impacted their ability to implement the intervention as planned. In addition, there were likely varying skill levels in the use of digital technology and in building supportive relationships with DAT users. In South Africa, research interns assisted TB focal nurses with implementation.

HCP engagement with the adherence platform was measured by several indicators. A higher percentage of weekday days with at least one visit to the platform per facility, indicates use of the platform was integrated into routine workflow, as seen in Ukraine at 76%. It is also possible that with more staff trained in Ukraine, there was a collective rather than individual action to make checking the platform routine practice, seen as key to professional behaviour change in complex healthcare settings (29). Daily reviewing of DAT data was indicated by 90% of HCPs in a meta-analysis of six studies (23), although this was a small sample self-reporting rather than verified through automated platform statistics as in this study. A cluster randomized trial in China concluded that HCPs did not use the electronic adherence data as intended, possibly due to frequent reports of pillboxes failing to log doses resulting from battery issues, although with a different platform and DAT (27). The large number of days with no facility staff accessing the adherence platform observed in South Africa and the Philippines could be explained by high workloads, or lack of motivation. A lack of trust in the adherence data, due to difficulty identifying those truly experiencing adherence issues may undermine staff motivation to use DATs (26), (30). Although qualitative data indicated use of DATs reduced workload, a finding also emphasized in other studies (28), it is possible that for HCPs already overburdened, and lacking digital skills, daily review of adherence may not have been feasible.

In both Philippines and Ukraine >50% of logins to the platform were from a smart phone rather than tablet or desktop, in contrast to the other three countries where most logins were via tablet. This may indicate that using a personal mobile phone was easier than using the facility tablet or desktop for some HCPs and aligns with a meta-analysis showing that 86% of HCP surveyed report assessing adherence using a mobile phone application (23). Accessing the platform using a mobile phone may also indicate that HCPs reviewed adherence outside of working hours, suggesting they may have been too busy during the workday. The existence of an electronic patient system as in the Philippines and Ukraine, in addition to the adherence platform may be a facilitator or a barrier to the intervention. Users of e-TB Manager in Ukraine gave high ratings to the registry’s ability to improve case management (31). TB staff were already accustomed to using digital systems, and had put in the time required to embed the behaviour change. However, they may also see it as additional work if they need to login to two separate systems for PwTB management, a finding from qualitative interviews with HCPs in Ukraine. Direct integration of adherence data into existing digital systems could mitigate this.

DAT coverage varied between countries, and the factors contributing to uptake of DATs were wide ranging. Staffing issues such as rotation in Tanzania or redeployment in Philippines, frequent power outages in South Africa, and healthcare reform as well as the war in Ukraine impacted coverage. Similar results were reported in a Ugandan trial with 52% coverage (3). Lower numbers of PwTB starting a DAT and variability over time underline the pragmatic nature of the trial where routine staff implement the intervention.

Manual dosing was an important part of the intervention, and provides a measure of HCP engagement with the intervention. The addition of manual doses to the adherence platform more than seven days after a missed dose was less than30% for both DAT types in Ethiopia, and greater than50% for both Philippines and Ukraine. HCPs would need to contact the participant to confirm if the dose was taken or not before adding a manual dose to the platform. Adding a manual dose within a short time of a missed dose may indicate a systematic approach to monitoring adherence, even if the capacity of the facility was low. Failure to add a manual dose within 1 week of a missed dose may indicate poor implementation or lack of engagement by HCP with the intervention, in addition to low capacity at the facility. It could also be due to inability to contact the PwTB. In Ukraine, the addition of manual dosing decreased significantly from pre-war to post start of the war, indicated by an increase of 7.8% to 30% of overall treatment days showing no information (32).

Overall, DAT engagement was high, measured by the percentage of digitally recorded doses. Label users recorded between 62% and 84% of doses by texting the toll-free number, conversely digital a study in Uganda where self-report of dose taking was done by calling 99DOTS was 58%, dropping further over the course of treatment (33). While a meta-analysis on DAT projects across ten countries showed that overall average adherence among PwTB with DS-TB varied between 80% and 90% (32). Engagement of pillbox users was higher than labels, with 82%-91% of doses digitally recorded. Sustained engagement offers a measure of reach of the intervention and high levels indicate users like DATs. A positive association between treatment satisfaction and medication adherence, and higher levels of treatment satisfaction among participants using a pillbox versus standard of care, was previously reported (34). Similarly, from the ASCENT trials, a study on feasibility and acceptability, showed that nearly all DAT users reported that using DATs motivated them to complete treatment (11). Furthermore, a recent scoping review suggest that video and pillbox DATs are generally acceptable with moderate to high levels of engagement (25).

The feeling of connection to their HCP was very high among participants in most countries, a finding which concurs to that of previous studies (35). It has been shown that better adherence to treatment may be facilitated by good PwTB-provider relationships (33), (36), (37) and the feeling of being care for may be enhanced by DATs. However, this was lower in Philippines, particularly for label users, (14). This is aligned with other results in Philippines, possibly indicating that HCPs had difficulty implementing the intervention as planned.

### Strengths and Limitations

The results of this study were based on data obtained from PwTB and HCPs in the intervention group taking part in five large pragmatic cluster randomised trials implemented across multiple healthcare facilities in five countries, increasing generalizability of findings. To our knowledge, this is the first process evaluation of a digital adherence technology intervention in tuberculosis, implemented in diverse contexts.

A strength of this study is the inclusion of sixteen indicators across intervention components. Measurement of many of the process indicators was based on data captured objectively by the adherence platform. A further strength of this research is its mixed method approach, data was obtained from multiple sources, including related sub-studies, which were integrated to form an overall result. The synthesis of both quantitative and qualitative evidence provide a comprehensive account of how DATs were implemented. In addition, the quantitative data is from a large sample, with results from 162 healthcare facilities in five countries, covering over 10,000 DAT participants and 552 HCPs.

A limitation of the study is that qualitative results are from secondary data collected for various sub-studies. The sub studies were designed to evaluate feasibility and acceptability of DATs through surveys and HCP and indepth participants interviews. Furthermore, the impact of contextual factors was not considered, such as technology or infrastructure downtime, or participants opting out of daily SMS reminders, this omission was due to lack of available data.

In conclusion, we observed variation in the level of fidelity to the intervention between countries. Daily utilization of the real-time data to monitor adherence, and subsequent performance of actions required for differentiated care, was impacted by the capacity of the health systems and staff working within it. Engagement with and acceptance of DATs is high and they facilitate enhanced connection with HCPs for engaged PwTB. DATs offer a valuable person-centred tool for supporting PwTB through treatment. Reach and fidelity of digital support interventions are influenced by factors at the health system, facility and individual levels, further work should focus on refining implementation strategies.

## Data Availability

The raw quantitative data supporting the conclusions of this article will be made available by the authors, without undue reservation. Selected quotes from qualitative interviews are in the manuscript.

## Notes

### Competing Interest Statement

The authors have declared no competing interest.

### Funding Statement

The author(s) declare financial support was received for the research, authorship, and publication of this article. This study was funded by Unitaid (grant agreement number: 2019–33-ASCENT) through the Adherence Support Coalition to End TB (ASCENT) project (https://www.digitaladherence.org/). The funding organization had no role in the study design, data collection, data analysis, data interpretation, or manuscript writing.

### Author Declarations

The study has been approved by the WHO Ethical Review Committee (0003296) and London School of Hygiene & Tropical Medicine Ethics Committee, UK (19135) following external peer review. Individual protocols have been reviewed and approved by relevant country-specific committees: Single Joint Research Ethics Board (Philippines SJREB 2019-57) Wits Human Research Ethics Committee (South Africa AUR2-1-268) Tanzania Medical Research Coordinating Committee (MRCC) at National Institute for Medical Research, Dar es Salaam (Tanzania NIMR/HQ/R.8a/Vol.IX/3431) and Ukraine Ethics Committee of Public Health Center of the MOH of Ukraine (Ukraine IRB 2019-33), under the overall ASCENT protocol. Written informed consents of TB patients for the main effectiveness study were collected from people with TB (PwTB) by the TB care providers at the intervention facilities. A waiver of consent was obtained to access TB register data. PwTB agreed to use the DAT and consent to have researchers use anonymised data collected to the ASCENT Adherence platform. Informed consents for the sub-studies were collected by research associates prior to the interviews. The individual-level data sets visible to research staff to monitor the study and conduct analysis are deidentified. All databases are maintained in password-protected systems. Where paper records exist, they are stored in the participating facilities in locked cabinets with access permitted to only relevant facility healthcare providers and research team members.

